# West African pre-pandemic cross-reactive antibody and cellular responses against SARS-CoV-2

**DOI:** 10.1101/2024.05.21.24307417

**Authors:** Bobby Brooke Herrera, Beth Chaplin, Souleymane MBoup, Adam Abdullahi, Michelle He, Sydney M. Fisher, Sulaimon Akanmu, Charlotte A Chang, Donald J Hamel, Ravindra K. Gupta, Phyllis J Kanki

**Author notes:** **Correspondence:** PK.

## Abstract

The COVID-19 pandemic had a severe impact globally, yet African populations exhibited unexpectedly lower rates of severe disease and mortality. We investigated the potential role of pre-existing immunity in shaping the epidemiology of COVID-19 in Africa. Leveraging paired plasma and peripheral blood mononuclear cells collected from Senegalese female sex workers prior to the COVID-19 pandemic, we observed substantial levels of pre-existing cross-reactive immunity to SARS-CoV-2, stemming from prior exposure to seasonal human coronaviruses (hCoVs). Our antibody analysis revealed a 23.5% (47/200) seroprevalence rate against SARS-CoV-2 nucleocapsid (N). Of these SARS-CoV-2 N-reactives, 85.1% (40/47), 44.7% (21/47), and 95.7% (45/47) showed antibody reactivity against hCoV-229E or hCoV-OC43 spike (S) and/or N or hCoV-HKU1 S. Our analysis of cellular responses also demonstrated cross-reactivity to SARS-CoV-2 with 82.2% (37/45) and 84.4% (38/45) showing IFN-γ responses against S and N, respectively. These findings suggest that prior hCoV exposure may induce cross-reactive adaptive immunity, potentially contributing to protection against COVID-19. A unique pre-pandemic subject had cross-reactive SARS-CoV-2 S antibodies with detectable neutralization and cellular responses. Our study provides unique insights into the dynamics of hCoV and SARS-CoV-2 immunity in West African populations and underscores the importance of understanding the role of pre-existing immunity in shaping COVID-19 outcomes globally.

## Introduction

The first COVID-19 cases were recorded on the African continent in March 2020, yet by the end of the pandemic in May 2023, Africa’s reported SARS-CoV-2 infections and deaths constituted only 2-4% of the global disease burden despite being home to ∼17% of the world’s population. Early projections of the COVID-19 pandemic’s impact on Africa predicted massive loss of life. Most African countries lacked adequate healthcare infrastructure, workforce, and equipped facilities to cope with a novel and highly infectious respiratory pathogen. However, other characteristics of most African countries suggested different outcomes from what was seen in resource-rich settings.^1^ Africa’s relatively young population age distribution with lower rates of co-existing morbidities, environmental factors, and pre-existing immunity from exposure to related viruses may explain the unexpected low rates of severe disease and mortality observed in Africa.^2^

In 2020, SARS-CoV-2 emerged as a new human coronavirus, even though worldwide populations were known to be regularly exposed to seasonal human coronaviruses (hCoVs) responsible for the “common cold,” including α-coronaviruses (hCoV-229E, hCoV-NL63,) and β-coronaviruses (hCoV-OC43, hCoV-HKU1). Although associated with mild illness, these hCoVs share genetic similarity to the SARS coronaviruses, which include SARS-CoV-1, SARS-CoV-2, and Middle East Respiratory Syndrome (MERS). hCoVs are globally endemic and estimated to be responsible for up to 15-30% of pre-pandemic annual respiratory infections, which are seasonal (e.g., fall and winter) and most prevalent in young children.

The widespread circulation of hCoVs with repeated exposure of human populations and shared sequence homology to SARS-CoV-2 led to the suggestion that hCoV immunologic memory could result in cross-reactive cellular and humoral responses that might in part explain the heterogeneity of COVID-19 presenting symptoms and pathogenic outcomes. While not providing sterilizing immunity, pre-existing hCoV immunity could reduce transmission and ameliorate symptoms of the related SARS-CoV-2. Therefore, research has been conducted to determine the role of pre-existing cross-reactive immunity to SARS-CoV-2. Multiple US and European studies described CD4+ T cell reactivity in SARS-CoV-2 unexposed individuals, attributed to pre-existing memory responses to hCoVs.^3–7^ In a UK household study, exposed contacts that remained PCR-negative showed significantly higher frequency of cross-reactive (p=.0139) and nucleocapsid (N)-specific IL-2 secreting memory T cells (p=.0355).^3^ Swadling et al. in a study of potentially abortive SARS-CoV-2 infection in UK healthcare workers (HCWs) described pre-existing T cell reactivity directed against the early transcribed replication-transcription complex (RTC).^4^ The RTC, consisting of RNA polymerase co-factor non-structural protein 7 (NSP7), RNA polymerase NSP12 and helicase NSP13, is expressed early in the viral life cycle and is highly conserved among members of *Coronaviridae*.^4^ Studies of pre-pandemic samples collected from Africa have also demonstrated cross-reactive T cell responses against several SARS-CoV-2 antigens.^5,6^ These studies have led to the hypothesis that prior hCoV infection might provide protective cross-reactive memory, perhaps more likely in younger individuals in whom infections with hCoVs are more prevalent and recent. As described by Lipsitch et al., pre-existing cross-reactive memory T cells could lower respiratory tract viral load thereby limiting the duration and severity of disease with potential to reduce viral spread.^7^

Pre-existing antibody studies have also described distinct humoral responses to SARS-CoV-2 associated with prior hCoV infection. Ng et al. described uninfected donors with IgG reactivity to conserved epitopes in the S2 subunit of the SARS-CoV-2 spike (S) protein distinct from de novo humoral responses in infected individuals that targeted both S1 and S2 subunits with concomitant IgM and IgA responses.^8^ Multiple studies conducted in the US and Europe have identified antibodies to hCoVs (OC43) that were associated with lower risk of severe COVID-19.^9, 10^

Over the course of the COVID-19 pandemic in Africa, studies based on SARS-CoV-2 antibody detection described high prevalence rates of infection, in contrast with surveillance data.^11^ Yet, even with anticipated underreporting, severe COVID-19 cases and mortality remained unexpectedly low.^12^ While multiple factors are likely to contribute to the capacity of individuals to generate cross-reactivity, it is possible that the order and composition of different infections may play a role in determining the efficacy of the immune response in preventing symptomatic or severe COVID-19. Currently, high rates of SARS-CoV-2 infection and COVID-19 vaccine exposure globally challenge the ability to investigate cross-reactive responses in individuals who have not been exposed to SARS-CoV-2. While our research group has reported on SARS-CoV-2 immune responses in Nigeria,^5^ in this study we leveraged paired plasma and peripheral blood monocular cell (PBMC) samples collected in Senegal, West Africa prior to the COVID-19 pandemic. Our objectives were to measure hCoV seroprevalence rates and to characterize the pre-existing cross-reactive antibody and cellular responses to SARS-CoV-2 in our pre-pandemic West African samples. The aim was to understand whether and how pre-existing hCoV immunity might confer protection against severe COVID-19 disease in Africa.

## Methods

### Human samples

The plasma and PBMC archive was developed from our multi-decade prospective cohort study of HIV-1 and HIV-2 in Senegalese female sex workers (FSWs), followed from 1986-2010.^13–15^ Self-identified FSW routinely visiting a health care clinic in Dakar provided blood samples to test for multiple sexually transmitted infections, including HIV-1 and HIV-2 and were examined clinically; excess blood samples were de-identified and archived with their corresponding clinical data. More than 80% of women sampled were HIV negative and most had paired plasma and liquid nitrogen preserved PBMCs with accompanying clinical and immunologic data at more than 2 timepoints. The viability and functional integrity of these cryopreserved PBMCs have been previously demonstrated in our studies of HIV and flavivirus T-cell immunity.^16–18^ The plasma and PBMC samples tested for this study were collected from 2004 to 2005. At the time of the original study, all women provided informed consent for the collection and archiving of samples and corresponding data; ethical clearance from the Harvard Institutional Review Board (IRB) and the research ethics committee at Chiekh Anta Diop University, Dakar, Senegal were obtained. All archived samples and corresponding data were anonymized, and as such this secondary analysis did not constitute human subject research by institutional guidelines and regulations which follow the Federal Policy for Protection of Human Subjects.

### Virion-based immunoblots

We screened plasma samples by immunoblot assay on virion preparations from SARS-CoV-2. Briefly, Vero E6 cells or susceptible cell line were infected with SARS-CoV-2 (Isolate USA-WA1/2020, BEI Resources NR-52281) or hCoVs (OC43, NL63, 229E, BEI Resources NR-5621, 470 and 52726, respectively) and propagated for five days. Supernatants were clarified at 10,000g for 20min at 4°C, precipitated with PEG-8000 and NaCl, and then resolved by sucrose gradient ultracentrifugation at 170,000g for 90min at 4°C. Viral pellets were lysed with complete NP40 buffer containing protease inhibitors. Viral lysates were added to nonreducing buffer (final concentrations of 2% SDS, 0·5 M Tris pH 6·8, 20% glycerol, 0·001% bromophenol blue) and subjected to 12% PAGE and Western blot analysis using patient serum (1:250) as primary antibody and anti-human IgG horseradish peroxidase (HRP) (1:2,000; ThermoFisher Scientific, Waltham, MA) as secondary antibody. Visualization was performed using Metal Enhanced DAB Substrate Kit (ThermoFisher Scientific, Waltham, MA) per the manufacturer’s instructions.

RecomLine SARS-CoV-2 IgG (Mikrogen)^19, 20^ is a CE-marked immunoblot assay which determines the IgG responses towards the recombinant N antigens of the seasonal hCoV 229E, OC43, NL63 and HKU-1 in parallel to SARS-CoV-2 N, receptor-binding domain (RBD), and S1 antigens.

### ELISPOT assay

ELISPOT assays were conducted as described previously^16, 21, 22^. In brief, 200,000 PBMCs per well were seeded in duplicate into 96-well plates coated with anti-human IFN-γ (Becton Dickinson (BD), Franklin Lakes, NJ). The cells were stimulated with fusion proteins consisting of a modified version of the *Bacillus anthracis* lethal factor (LFn) and SARS-^21^CoV-2 S or N ((Mir Biosciences, Inc., Dunellen, NJ) and either a negative (LFn alone, Mir Biosciences, Inc., Dunellen, NJ) or positive (PHA, ThermoFisher, Waltham, MA) control at 37°C with 5% CO_2_ for 24 hours. LFn has been shown to transport protein cargoes into the cell cytosol for MHC Class I and II processing, inducing T cell responses.^16, 21, 22^ After incubation, plates were washed followed by incubation for 2 hours at room temperature with a secondary antibody (BD, Franklin Lakes, NJ). Subsequently, plates were washed and incubated with streptavidin (BD, Franklin Lakes, NJ) for 1 hour at room temperature. Finally, plates were washed and incubated with substrate (BD, Franklin Lakes, NJ) for 30 minutes. IFN-γ spot forming cells (SFC) were counted using the ImmunoSpot S6 Ultra M2 Analyzer ^16, 21, 22^ (ImmunoSpot, Shaker Heights, OH)

#### Virus neutralization titer analyses

Pseudotype virus neutralization assay was performed on Hela-ACE2 cells using SARS-CoV-2 spike pseudotype virus (PV) expressing luciferase as previously described.^23, 24^ Briefly, dried plasma spots were eluted and heat inactivated at 54°C for 1 hour,^25^ and incubated with PVs at 37°C for 1 hour prior to addition of Hela-ACE2 cells. The plasma dilution/virus mix was incubated for 48 hours in a 5% CO_2_ environment at 37°C, and luminescence was measured using the Bright-Glo Luciferase assay system (Promega UK, United Kingdom). Neutralization was calculated relative to the virus and cell only controls. Data was analyzed in GraphPad Prism v9.3.1 where 50% neutralization (ID_50_) values were calculated and the limit of detection for neutralization was set at ID_50_ of 20 units.

#### Statistics

Bivariate analyses explored associations between select demographic characteristics of the women with available data and SARS-CoV-2 N-reactive immunoblot results. Demographic characteristics included: year of sampling, age at sampling, neighborhood, ethnicity, country of origin, marital status, religion, education, and total children. The chi-square test was used to obtain p-values to detect statistically significant associations, and Fisher’s exact test for categories with observed frequencies ≤5.

## Results

### Pre-pandemic cross-reactive SARS-CoV-2 antibody responses and hCoV seroprevalence rates

Pre-pandemic plasma samples (n=200) from the Senegalese FSW cohort were analyzed for the presence of antibodies directed against SARS-CoV-2 S and/or N. 152 of 200 (76%) were negative, while 47 (23.5%) showed reactivity to SARS-CoV-2 N, suggestive of prior exposure to hCoVs (Fig. 1, Fig. 2A). To determine hCoV seroprevalence rates, the 47 samples reactive to SARS-CoV-2 N-only were analyzed for antibodies directed against hCoV-229E S and N, hCoV-OC43 S and N, and hCoV-HKU1 S. Of the samples tested, 37 (78.7%) samples showed reactivity to hCoV-229E S±N, 7 (14.9%) to hCoV-OC43 S±N, and 45 (95.7%) to hCoV-HKU1 S (Fig. 1, Fig. 2B-D). Of the 37 hCoV-229E S±N samples, 30 (81.1%) were reactive to S+N, while 7 (18.9%) were reactive to S-only. Of the 7 hCoV-OC43 S±N samples, 6 (85.7%) were reactive to S+N, while 1 (14.3%) was reactive to S-only. Of note, 36 out of the 47 samples (76.6%) were reactive to both hCoV-229E S and hCoV-HKU1 S (Supplementary Fig. 1). Only 6 (12.8%) were reactive to S of hCoV-229E, hCoV-OC43, and hCoV-HKU1 (Supplementary Figs. 2-3).

**Fig. 1.**
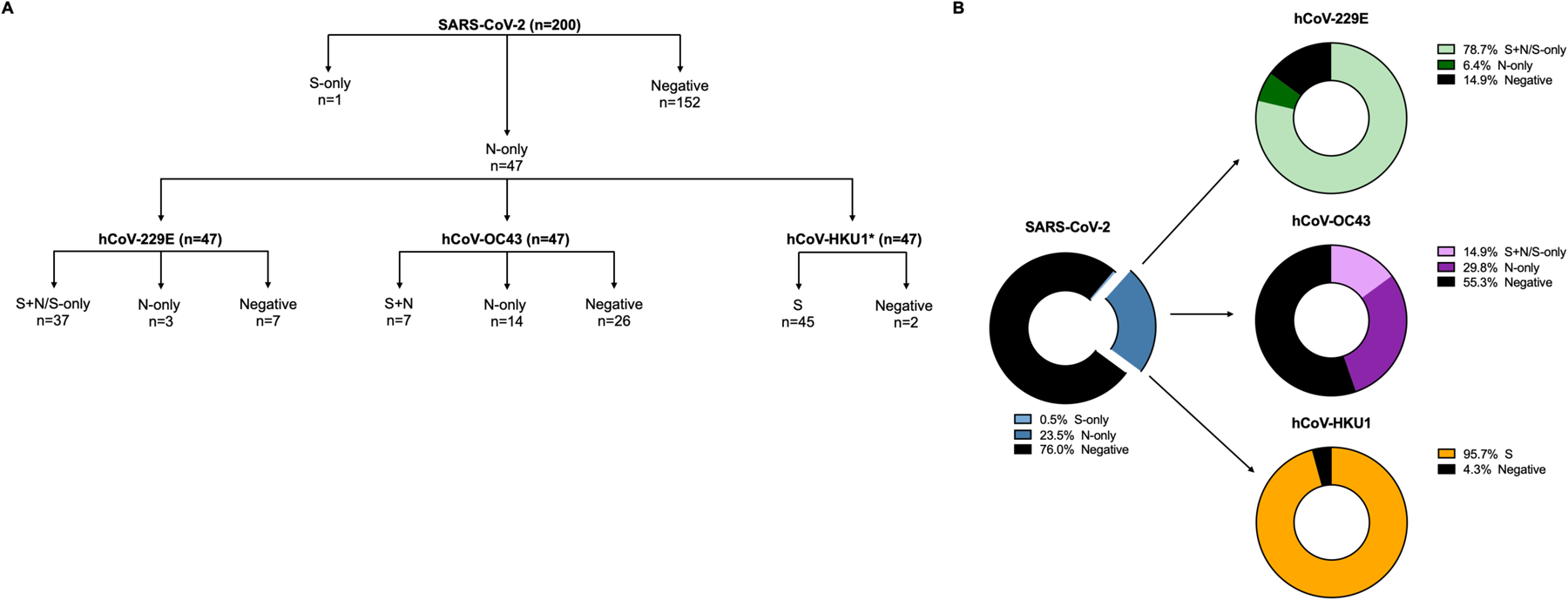
SARS-CoV-2 and hCoVs seroprevalence rates. A) Schematic representation of serology workflow. *, Samples were analyzed against hCoV-HKU1 S only. B) 200 samples were initially tested against SARS-CoV-2 S and N; 47 samples had antibodies against SARS-CoV-2 N-only (blue donut chart). These 47 samples were then tested against hCoV-229E S and N (green donut chart), hCoV-OC43 S and N (purple donut chart), and hCoV-HKU1 S (yellow donut chart).

**Fig. 2.**
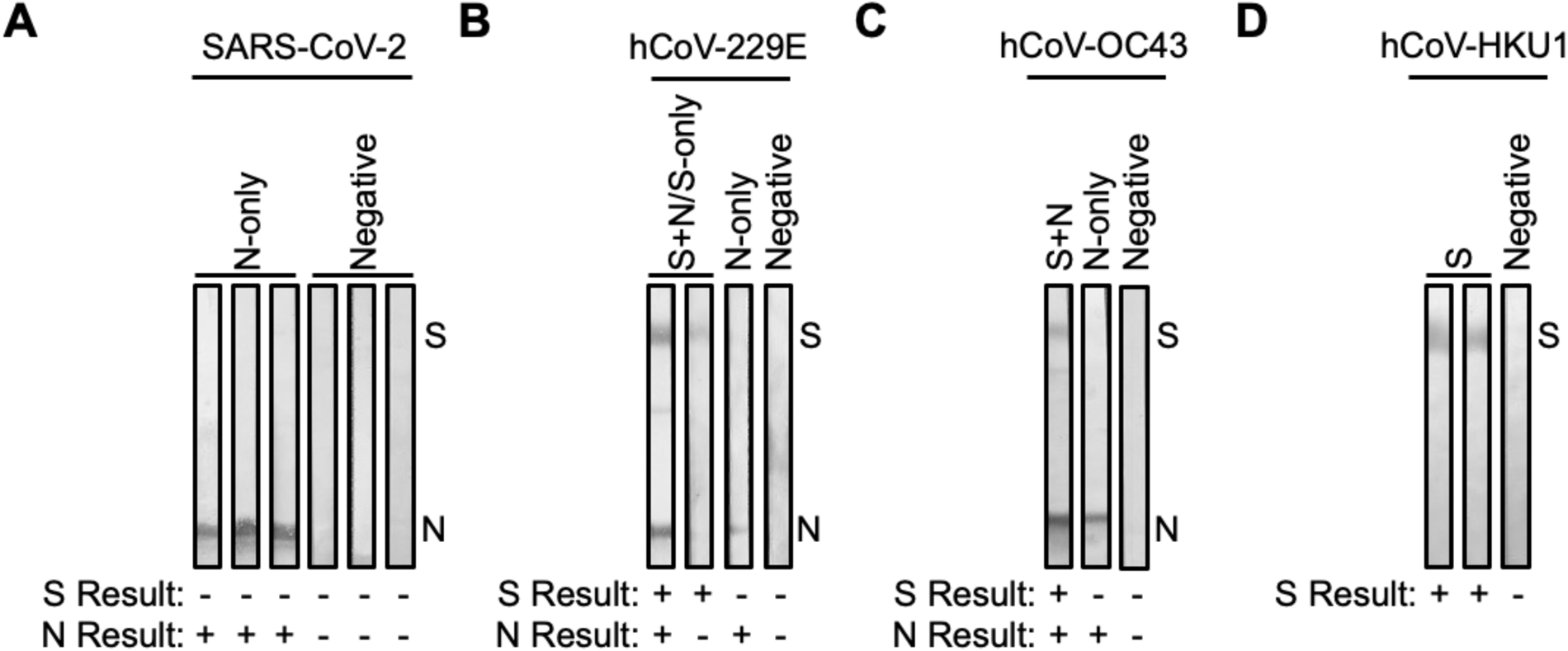
SARS-CoV-2 and hCoV antibody cross-reactive in samples collected pre-pandemic. Plasma collected from FSWs prior to the COVID-19 pandemic were subjected to immunoblot analysis. Representative images of immunoblots for samples that had antibodies for A) SARS-CoV-2 N-only or were negative, B) hCoV-229E S+N/S-only, N-only, or were negative, C) hCoV-OC43 S+N, N-only, or were negative, and D) hCoV-HKU1 S or were negative.

### Risk factor analysis

Bivariate analysis of risk factors associated with SARS-CoV-2 N antibody reactivity included: year of sampling, age at sampling, neighborhood, ethnicity, country of origin, marital status, religion, education, and total children. There was a potential association between SARS-CoV-2 N-reactive and age >44 years (p=0.078) and with having ≥3 children (p=0.044) (Table 1). Older age had a strong positive correlation with having more children.

**Table 1.** Associations between baseline characteristics and SARS-CoV-2 nucleocapsid (N) antibody reactivity (N=200^1^)

|  | N reactive |  | N non-reactive |  | Chi-square* p-value |
| --- | --- | --- | --- | --- | --- |
|  | No. | % | No. | % |  |
| Year of sampling |  |  |  |  |  |
| 2004 | 74 | 81.3% | 17 | 18.7% | 0.142 |
| 2005 | 79 | 72.5% | 30 | 27.5% |  |
| Age at sampling |  |  |  |  |  |
| ≤44 years | 92 | 76.0% | 29 | 24.0% | 0.078 |
| >44 years | 22 | 61.1% | 14 | 38.9% |  |
| Neighborhood |  |  |  |  |  |
| Dakar | 82 | 75.2% | 27 | 24.8% | 0.582 |
| Outside Dakar | 37 | 71.2% | 15 | 28.8% |  |
| Ethnicity |  |  |  |  |  |
| Wolof | 47 | 78.3% | 13 | 21.7% | 0.188 |
| Other | 68 | 68.7% | 31 | 31.3% |  |
| Country of origin |  |  |  |  |  |
| Senegal | 106 | 73.1% | 39 | 26.9% | 0.535* |
| Other | 9 | 64.3% | 5 | 35.7% |  |
| Marital status |  |  |  |  |  |
| Single | 34 | 77.3% | 10 | 22.7% | 0.578* |
| Divorced | 64 | 71.9% | 25 | 28.1% |  |
| Widowed | 7 | 63.6% | 4 | 36.4% |  |
| Religion |  |  |  |  |  |
| Muslim | 94 | 73.4% | 34 | 26.6% | 0.767* |
| Christian | 11 | 68.8% | 5 | 31.3% |  |
| Education |  |  |  |  |  |
| None | 43 | 69.4% | 19 | 30.6% | 0.428 |
| 1-6 years | 30 | 81.1% | 7 | 18.9% |  |
| 7-14 years | 30 | 71.4% | 12 | 28.6% |  |
| Total children |  |  |  |  |  |
| 0-2 | 70 | 78.7% | 19 | 21.3% | 0.044 |
| ≥3 | 45 | 64.3% | 25 | 35.7% |  |

<sup>1</sup> 200 samples tested: 47 SARS-CoV-2 N positive, 153 SARS-CoV-2 N negative. 159 had baseline record.
\*Fisher's exact test was used for variables with cells with observed frequencies ≤5.

### Pre-pandemic cross-reactive T-cell responses to SARS-CoV-2

Of the 47 pre-pandemic samples that showed antibody reactivity to SARS-CoV-2 N-only and also antibody reactivity to hCoV S+N/S, 45 paired PBMCs were available, along with 21 paired PBMCs from negative controls, and were analyzed for cross-reactive IFN-γ cellular responses against SARS-CoV-2 S and N. Among SARS-CoV-2 N-reactives, 37 (82.2%) showed IFN-γ reactivity against SARS-CoV-2 S (Fig. 3A-B) while 9 (19.6%) were negative. Similarly, 38 (84.4%) showed IFN-γ reactivity against SARS-CoV-2 N (Fig. 3C-D) while 8 (17.4%) were negative. Among the 21 antibody negative controls, 3 (14.3%) demonstrated weak IFN-γ cellular reactivity against both SARS-CoV-2 S and N.

**Fig. 3.**
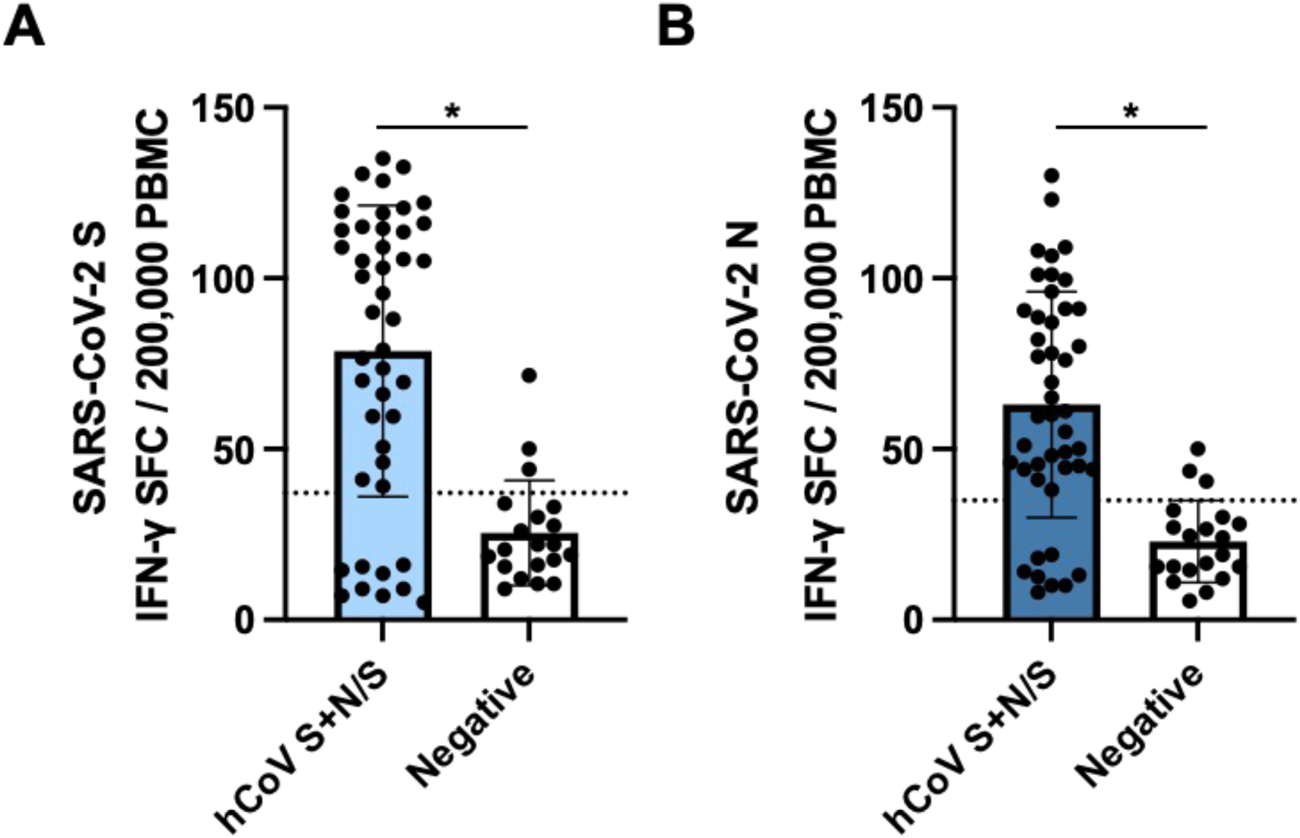
Cross-reactive SARS-CoV-2 cellular responses in samples collected pre-pandemic. PBMCs collected from FSWs prior to the COVID-19 pandemic were stimulated with A) LFn-SARS-CoV-2 S or B) LFn-SARS-CoV-2 N and IFN-γ responses were detected by ELISPOT. SFC, spot forming cells. Black dotted lines, LFn-SARS-CoV-2 S and N cutoffs.

### Evidence for pre-pandemic SARS-CoV-2 variant

In our initial serology, a single sample collected from a FSW in November 2004 demonstrated strong antibody reactivity to both SARS-CoV-2 S and N (Fig. 1A). Longitudinal antibody analysis of 6 samples collected from this individual between 1997 to 2004 was conducted by SARS-CoV-2 immunoblot as well as the Mikrogen assay that detected antibodies against the individual antigens of SARS-CoV-2 (S, RBD, and N) and hCoVs (HKU1, OC43, NL63, and 229E N). Samples collected in 1997 and 2001 showed antibody reactivity to SARS-CoV-2 N-only by immunoblot with weak antibodies against hCoVs, particularly in the 1997 sample (Fig. 4A-B). Seroconversion to SARS-CoV-2 S (2003) and S and RBD (May to November 2004) was observed by both immunoblot and the Mikrogen assay, accompanied with stronger reactivity to hCoV-HKU1 N and hCoV-OC43 N. This seroprofile is unique from our other pre-pandemic serology and may indicate exposure to a novel variant – one more closely related to SARS-CoV-2. Additionally, longitudinal cellular responses were analyzed between 1997 and 2004. In 1997, this individual only had cellular responses against SARS-CoV-2 N. By 2001, this individual had cellular responses against both SARS-CoV-2 S and N that persisted throughout 2004 (Fig. 4C). Sequential analysis of SARS-CoV-2 neutralization revealed no detectable neutralization response against SARS-COV-2 wild-type (Wu-1 614G) in samples collected in 1997 and 2001 but showed detectable neutralization response in samples collected in October 2003 (ID_50_ titer=281.3) with waning responses observed in May 2004 (ID_50_ titer=195.6); August 2004 (ID_50_ titer=121.1) and November 2004 (ID_50_ titer=43.3). The observation of detectable responses coincided with seroconversion of S and RBD antibodies reflecting exposure to hCoVs with possible cross-reactive potential due to sequence homogeneity to a SARS-CoV-2 variant. Clinical examination data collected as part of the prospective HIV clinical protocol did not indicate reporting of symptoms or signs of respiratory disease, but the patient was noted to have lost significant (>10%) weight between 2001 to 2003, during which S antibody seroconversion occurred. The patient was also HIV-positive, 43 years of age in 2003, and had 3 living children.

**Fig. 4.**
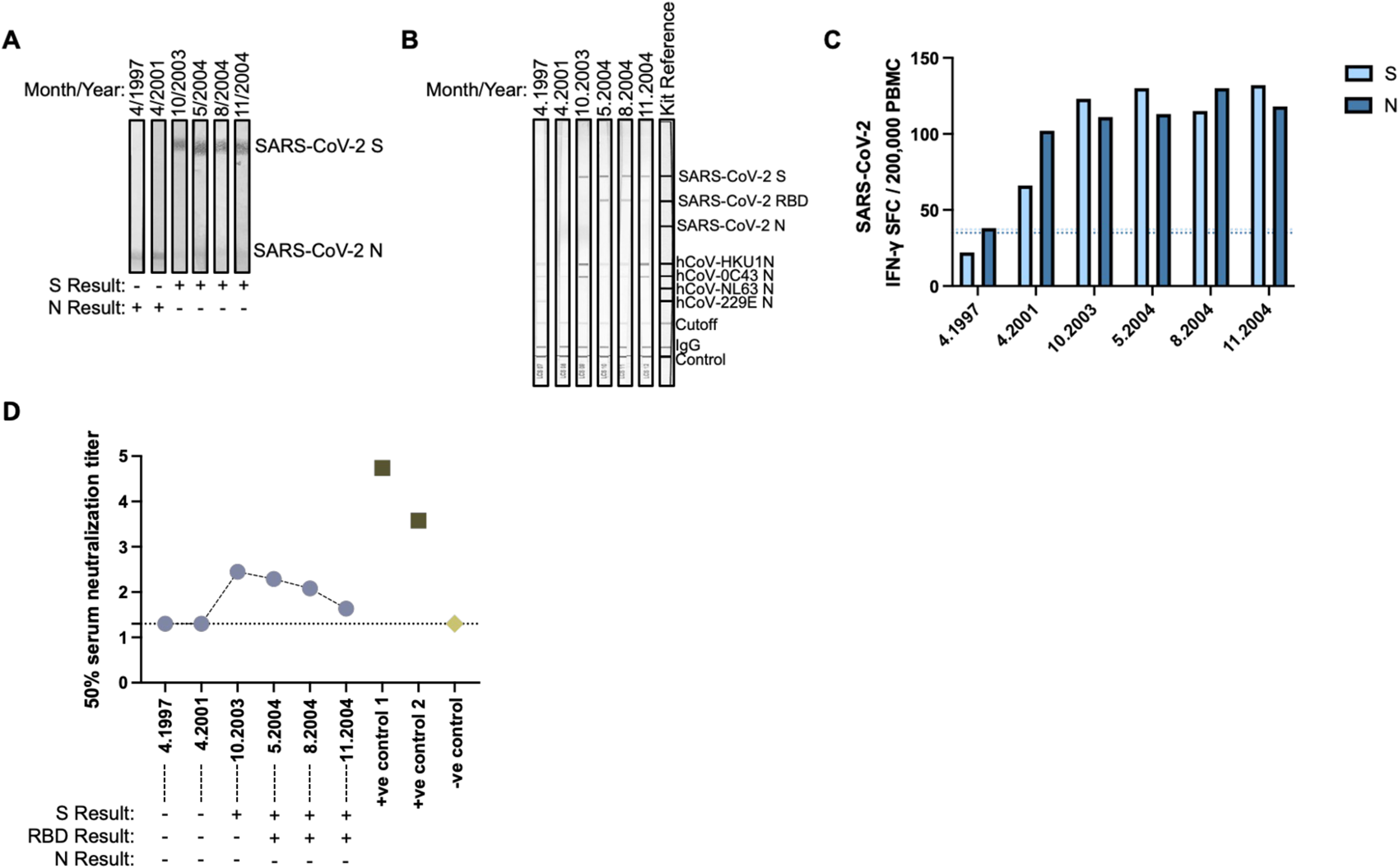
Evidence for pre-pandemic SARS-CoV-2 variant. Longitudinally collected plasma samples collected from a FSW between 1997 and 2004 were subjected to A) SARS-CoV-2 immunoblots or the commercial B) Mikrogen assay. C) Paired PBMCs samples were stimulated with LFn-SARS-CoV-2 S or LFn-SARS-CoV-2 N and IFN-γ responses were detected by ELISPOT. SFC, spot forming cells. Light blue dotted line, LFn-SARS-CoV-2 S cutoff. Dark blue dotted line, LFn-SARS-CoV-2 N cutoff. D) Plasma neutralization (Log10) of pseudotype virus against SARS COV-2 Wild type (Wu-1) in one individual with longitudinal samples collected between 1997 and 2004 including two positive controls from SARS CoV-2 infected and vaccinated adults in 2021 and pre-pandemic human sera as negative control. Results displayed under the x-axis correspond to their antibody reactivity via immublot assays.

## Discussion

In early 2021, we studied the immune responses in Nigerian healthcare workers (HCWs) at Lagos University Teaching Hospital (n=134) and in Oxford/AstraZeneca COVID-19 vaccine recipients from the general population (n=116) across five local government areas (LGAs) in Lagos state.^5^ Antibodies directed to only SARS-CoV-2 N in the absence of S antibody reactivity were detected in 9.7% (13/134) of HCWs and 15.5% (18/116) of the general population. This antibody profile directed to the SARS-CoV-2 N alone was suggestive of pre-existing cross-reactive immunity stemming from prior hCoV infection. To investigate this possibility further, in this study we tested archived plasma samples collected before the COVID-19 pandemic from our FSW cohort in Dakar, Senegal (2004-2005), and observed 23.5% (47/200) of individuals with pre-existing immunity to SARS-CoV-2 N-only (Fig. 1). Further testing demonstrated prior infections with the seasonal hCoVs, providing evidence that the SARS-CoV-2 N-only reactivity resulted from cross-reactivity from prior hCoV exposure (Fig. 1-2). The coronavirus N protein is more conserved than the S protein. As such, previously hCoV exposed individuals can demonstrate cross-reactive N antibodies without cross-reactive S antibodies (Figs. 1-2).^5, 26^

In one of the few studies conducted on samples from Africa prior to the COVID-19 pandemic, Tso et al. reported higher reactivity to hCoV N and S antigens with significantly higher rates of cross-reactivity in Zambia (14.1%) and Tanzania (19%) compared to the US (2.4%).^27^ In a multi-national study, Pedersen et al. reported higher SARS-CoV-2 N reactivity in Gabon (20/116,17.2%) compared to reactivity in Canada, Denmark, and Brazil. In samples from Senegal, largely from subjects under 17 years of age, high antibody cross-reactivity to both S and N antigens of SARS-CoV-2 was observed; these samples failed to demonstrate in vitro neutralization.^26^ Similar to this Senegal study, our study also detected pre-pandemic antibody cross-reactivity to SARS-CoV-2, but we concurrently detected cellular cross-reactivity to support the antibody results.

While most investigators study T cell responses to specific antigens using overlapping peptides, we have used full-length antigens or large peptides fused to a modified version of the *Bacillus anthracis* lethal factor (LFn). LFn has been shown to act as a molecular shuttle, transporting antigens of interest into the cytosol for MHC Class I and Class II processing for CD4+ and CD8+ T cell presentation. In the past, we have used LFn to study T cell responses against several viruses, including HIV, Ebola, Zika, dengue, and SARS-CoV-2. ^22–24,18^ In our SARS-CoV-2 study of Nigerian HCWs and individuals from the general population, among individuals with SARS-CoV-2 N-only antibodies prior to vaccination, suggestive of prior exposure to hCoV, 81.8% (9/11) had T cell responses to SARS-CoV-2 N. These results provided further evidence that individuals who were previously exposed to hCoVs could have cross-reactive cellular responses against SARS-CoV-2.

Cross-reactive T cell responses play a protective role in protection against COVID-19. Cellular responses were shown to map to cross-reactive recognition of SARS-CoV-2 by T cells induced by hCoVs.^28–31^ SARS-CoV-2-specific T cells were also able to cross-react with SARS-CoV-1 and MERS-CoV.^32–35^ Recently, Tarke et al., demonstrated that pre-pandemic samples collected from healthy adults in the US with confirmed previous exposure to hCoV-NL63 and -OC43 had significant cross-reactive T cell responses to SARS-CoV-2, particularly to conserved regions of SARS-CoV-2 S, N, nsp3, and nsp12.^36^ Similarly, Namuniina et al. showed that in pre-pandemic samples collected in Uganda, a majority of subjects had both CD4+ and/or CD8+ T cell responses against SARS-CoV-2 S and non-S peptide pools.^6^ This study however did not determine whether pre-existing immunity based on previous exposure to hCoV was responsible for the cross-reactive cellular responses.

In the current study using pre-pandemic samples collected from Senegalese FSWs, we show evidence of cellular cross-reactivity to SARS-CoV-2 with 82.2% (37/45) and 84.4% (38/45) of PBMC samples matched to SARS-CoV-2 N antibody reactive plasma samples demonstrating IFN-γ responses against S and N, respectively (Fig. 3). Although these N antibody reactive samples showed no S antibody reactivity, the majority demonstrated cross-reactive cellular responses to both SARS-CoV-2 S and N, suggesting that cellular responses against this less conserved region may be more sustained over time than humoral responses. Among the SARS-CoV-2 negative controls, a majority (18/21, 85.7%) did not have cellular responses. However, three control individuals showed weak cellular responses against SARS-CoV-2 S and N. This again may reflect the more temporal nature of humoral immunity compared with the longevity of cellular immunity. Additionally, the individual that initially tested antibody positive for SARS-CoV-2 S-only also had sustained cross-reactive cellular responses to SARS-CoV-2 S and/or N from 1997 to 2004 (Fig. 4C) with evidence of sustained albeit waning neutralization responses from 2003 and 2004 (Fig. 4D).

It is unknown whether these cross-reactive cellular responses from prior hCoV infection were sustained at levels that would impact the pathogenesis of SARS-CoV-2 during the 2020 pandemic. Immunologic memory to antigenically related infections can result in both positive and negative patient outcomes. As described in influenza, the induction of antibody responses to new virus infection can “backboost” responses to preceding heterologous viruses.^37^ In contrast, prior heterologous responses with reduced functionality may misdirect new responses resulting in negative clinical outcomes, i.e. “original antigenic sin”^38, 39^, particularly in antigenically shifting viruses such as SARS COV-2. Our results show significant prior hCoV infection with demonstrable antibodies to SARS-CoV-2 N and cellular responses to S and N, which when taken in consideration with low rates of severe COVID-19 disease and mortality in Africa, suggest through indirect association the possibility that prior hCoV infection provides cross-reactive protective immunity in SARS-CoV-2 infection.

In our risk factor analysis, we saw potential higher risk of SARS-CoV-2 N reactivity, suggesting previous hCoV infection, in women who were >44 years of age and who had ≥3 children. This might be explained by older women having had more children, and therefore increased exposure to respiratory illnesses from young and school-aged children amongst whom susceptibility to infection and transmission may be highest.

Our study of pre-pandemic immunologic reactivity to SARS-CoV-2 in Senegal is unique in its assessment of both antibody and cellular responses. However, our study has limitations. We were unable to evaluate the cellular immune responses to hCoVs to verify the proposed source of SARS CoV-2 cross-reactive IFN-γ cellular responses. Our assessment of primary hCoV infection was based on serologic reactivity to hCoV S, which may not be diagnostic of hCoV infection or correlate with hCoV N reactivity or cross-reactivity to SARS-CoV-2 N. We were unable to directly test our hypothesis by demonstrating that a significant fraction of the population possessed antibody and cellular cross-reactive immune responses to SARS-CoV-2 at the time of the pandemic. Also, our study analyzed samples from only adult females, who may have different immune responses than males and children.

Altogether, our study uniquely analyzed both antibody and cellular responses in West Africans, demonstrating that pre-existing immunity against hCoVs can induce cross-reactive T cell responses against SARS-CoV-2. The concurrent measurement of cross-reactive immune responses from both the antibody and cellular arms of the adaptive response provides additional support to the hypothesis that pre-existing hCoV adaptive immunity might impact SARS CoV-2 pathogenesis. These data provide valuable insights toward a clearer understanding of the dynamics of SARS-CoV-2 disease progression and protection in the West African setting.

## Data Availability

All data produced in the present work are available upon reasonable request to the authors.

---

**Supplementary Fig. 1.**
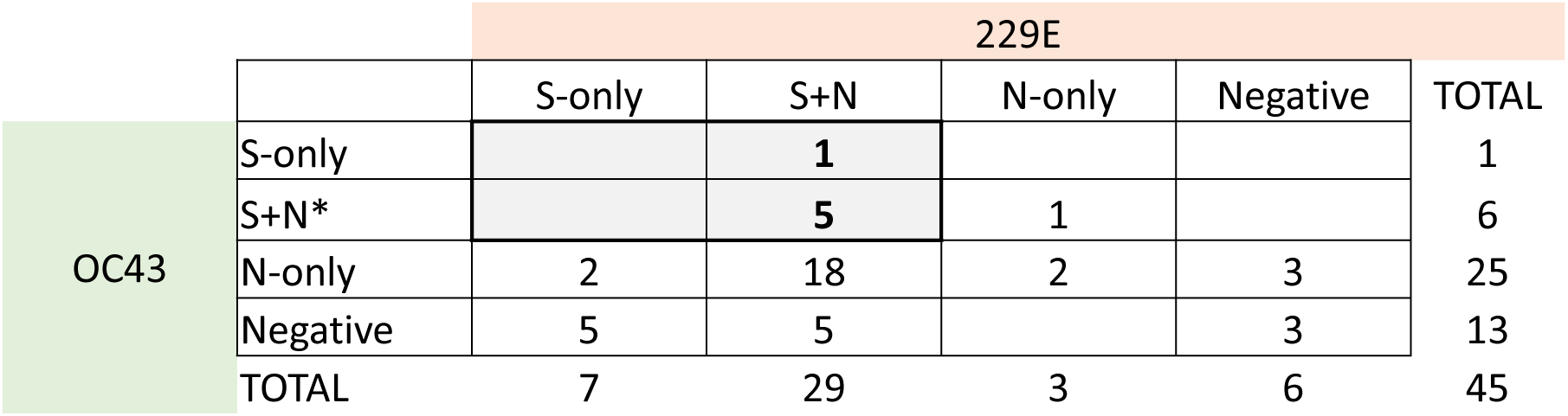
Overlap in reactivity between hCoV-229E and hCoV-OC43

**Supplementary Fig. 2.**
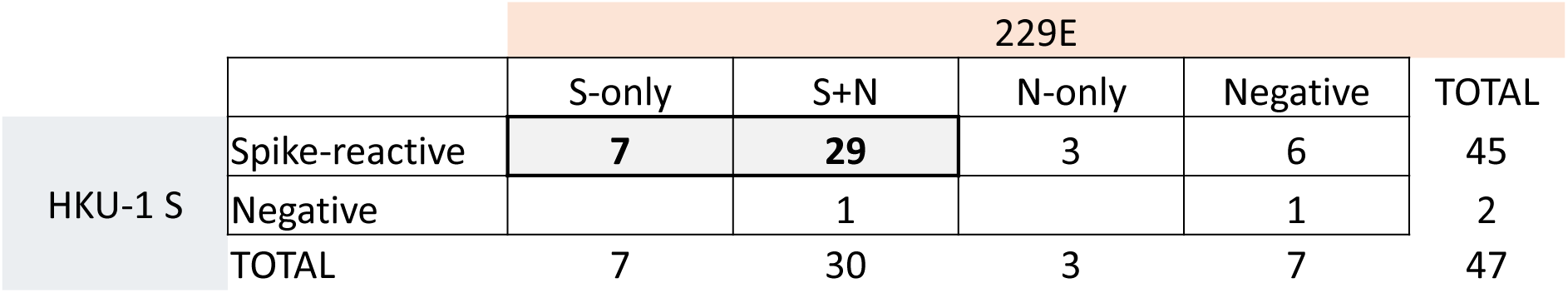
Overlap in reactivity between hCoV-229E S and hCoV-HKU1 S.

**Supplementary Fig. 3.**
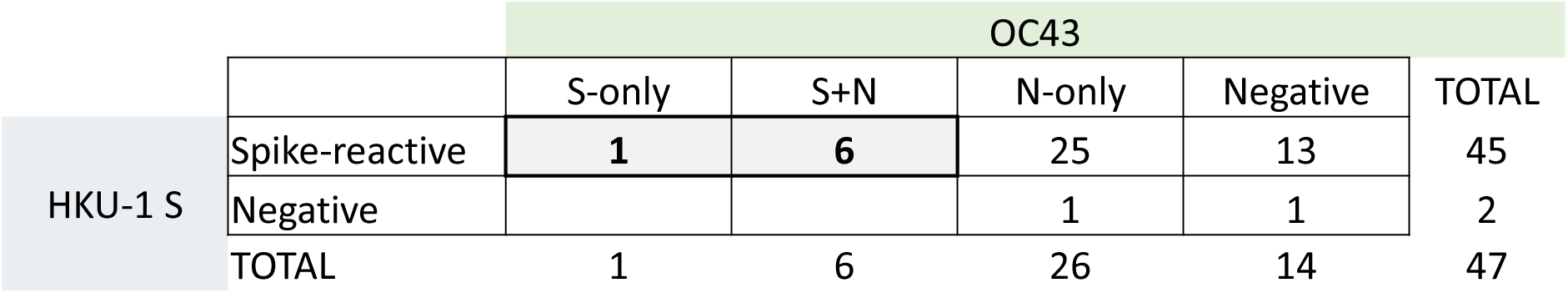
Overlap in reactivity between hCoV-OC43 S and hCoV-HKU1 S.

